# Viral genome sequencing places White House COVID-19 outbreak into phylogenetic context

**DOI:** 10.1101/2020.10.31.20223925

**Authors:** Trevor Bedford, Jennifer K. Logue, Peter D. Han, Caitlin R. Wolf, Chris D. Frazar, Benjamin Pelle, Erica Ryke, James Hadfield, Jover Lee, Mark J. Rieder, Deborah A. Nickerson, Christina M. Lockwood, Lea M. Starita, Helen Y. Chu, Jay Shendure

**Affiliations:** Vaccine and Infectious Disease Division, Fred Hutchinson Cancer Research Center, Seattle, WA, USA; Brotman Baty Institute for Precision Medicine, Seattle, WA, USA; Department of Genome Sciences, University of Washington, Seattle, WA, USA; Department of Medicine, Division of Allergy and Infectious Diseases, University of Washington, Seattle, WA, USA; Department of Laboratory Medicine and Pathology, University of Washington, Seattle, WA, USA; Howard Hughes Medical Institute, Seattle, WA, USA

## Abstract

In October 2020, an outbreak of at least 50 COVID-19 cases was reported surrounding individuals employed at or visiting the White House. Here, we applied genomic epidemiology to investigate the origins of this outbreak. We enrolled two individuals with exposures linked to the White House COVID-19 outbreak into an IRB-approved research study and sequenced their SARS-CoV-2 infections. We find these viral sequences are identical to each other, but are distinct from over 190,000 publicly available SARS-CoV-2 genomes. These genomes fall as part of a lineage circulating in the USA since April or May 2020 and detected in Virginia and Michigan. Looking forwards, sequencing of additional community SARS-CoV-2 infections collected in the USA prior to October 2020 may shed further light on its geographic ancestry. In sequencing of SARS-CoV-2 infections collected after October 2020, it may be possible to identify infections that likely descend from the White House COVID-19 outbreak.

## Introduction

After its emergence in late 2019, COVID-19 spread throughout the world, resulting globally in over 42 million cases and over 1.1 million deaths as of October 25, 2020 [1]. In the United States alone, there have been over 8.9 million confirmed cases and over 225,000 deaths reported [2]. COVID-19 has been repeatedly associated with localized outbreaks surrounding social settings like weddings and bars [3] as well as workplaces [4], including so-called “superspreader” events. Social events and workplaces, among other exposure settings, are thought to be driving ongoing transmission in the United States [5]. In October 2020, an outbreak of COVID-19 was reported surrounding individuals employed at the White House, individuals attending an event at the White House Rose Garden announcing Amy Coney Barrett’s nomination to the Supreme Court and individuals attending other events between September 26 and September 30, 2020 [6]. As of October 30, 2020, there are at least 50 individuals who have publicly announced cases of COVID-19 linked to this outbreak [7]. The origins of the White House outbreak have been characterized as “unknowable” [8].

Due to long incubation times and a spectrum of disease severity, contact tracing of COVID-19 spread is challenging. However, genomic sequencing of the SARS-CoV-2 virus causing individuals’ infections offers an alternative avenue for investigating SARS-CoV-2 transmission and spread [9–11]. This technology enables genomic epidemiology where genetic relationships among sequenced samples are used to make inferences about how infections are epidemiologically related. Because SARS-CoV-2 mutates approximately once every two weeks along a transmission chain [9,12], it is possible to use patterns of shared mutations to group viruses and discover transmission relationships.

We applied genomic epidemiology to shed light on the origins of the White House outbreak. We enrolled two individuals with exposures linked to the White House COVID-19 outbreak into an IRB-approved research study, collected nasal swabs and sequenced the SARS-CoV-2 virus from these specimens. These two individuals reported no direct contact with each other in the days preceding their COVID-19 diagnoses and are independently linked to the White House COVID-19 outbreak. We refer to these infections as WH1 and WH2. We report here on the genetic relationships between WH1, WH2 and publicly available SARS-CoV-2 genomic data.

## Results

Samples WH1 and WH2 were each obtained as an anterior nasal swab from individuals who had previously tested positive for SARS-CoV-2 shortly after attending event(s) associated with the White House COVID-19 outbreak. After confirmation of the positive test (Ct = 25 for Orf1b and S for WH1 and Ct = 25 for Orf1 and S for WH2), we performed genome sequencing on both samples either using a hybrid capture approach. This sequencing resulted in 550X average depth-of-coverage of WH1, yielding a consensus genome of 29,857 resolved bases or 99.8% of reference, and 5575X average depth-of-coverage of WH2, yielding a consensus genome of 29,858 resolved bases or 99.8% of reference. WH1 and WH2 are identical to each other at the consensus level. They possess 14 mutations relative to the reference strain Wuhan/Hu-1/2019 (241T, 1059T, 1977G, 3037T, 7936T, 14250T, 14408T, 16260T, 18417C, 19524T, 20402T, 23403G, 25563T, 28821A) shown in **Figure 1**. Of 195,737 publicly available viral sequences, only WH1 and WH2 share all 14 of these mutations. Genetically identical sequences for WH1 and WH2 support a close epi linkage between these infections and thus support independent infections from the same outbreak.

**Figure 1.**
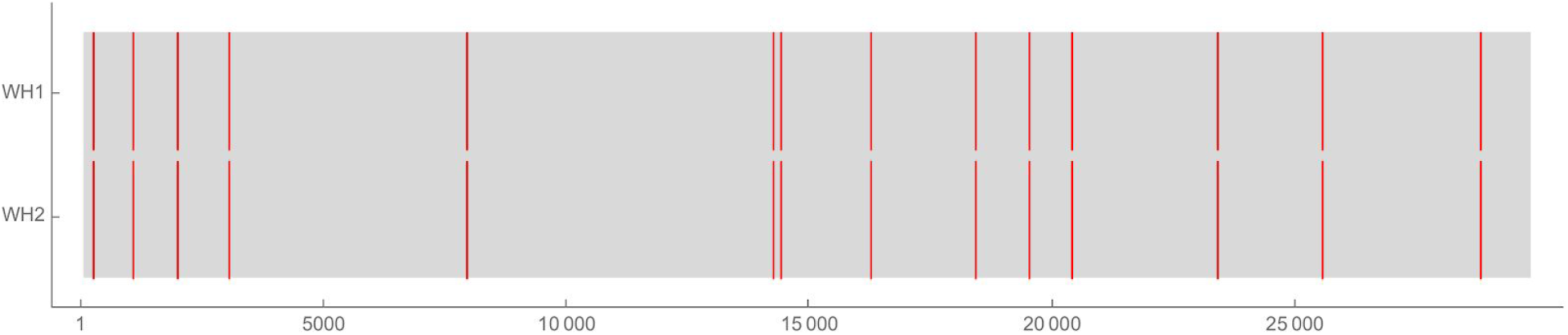
Rugplot showing coverage in gray and mutations relative to reference in red in WH1 and WH2. There are 14 distinct mutations shared by WH1 and WH2.

These mutations place WH1 and WH2 squarely within circulating genetic diversity in the United States (**Fig. 2**). WH1 and WH2 belong to Nextstrain clade 20C, which is the clade that predominates in the USA epidemic [13] and additionally belong to Pangolin lineage B.1.26 [14]. Pangolin lineage B.1.26 is demarcated by mutations C16260T and C28821A and contains viruses sampled from the USA, Canada and New Zealand (cov-lineages.org/lineages/lineage_B.1.26.html). The New Zealand samples comprise a 98-case cluster which was closed in June 2020 and whose source is epidemiologically linked to the USA [15].

**Figure 2.**
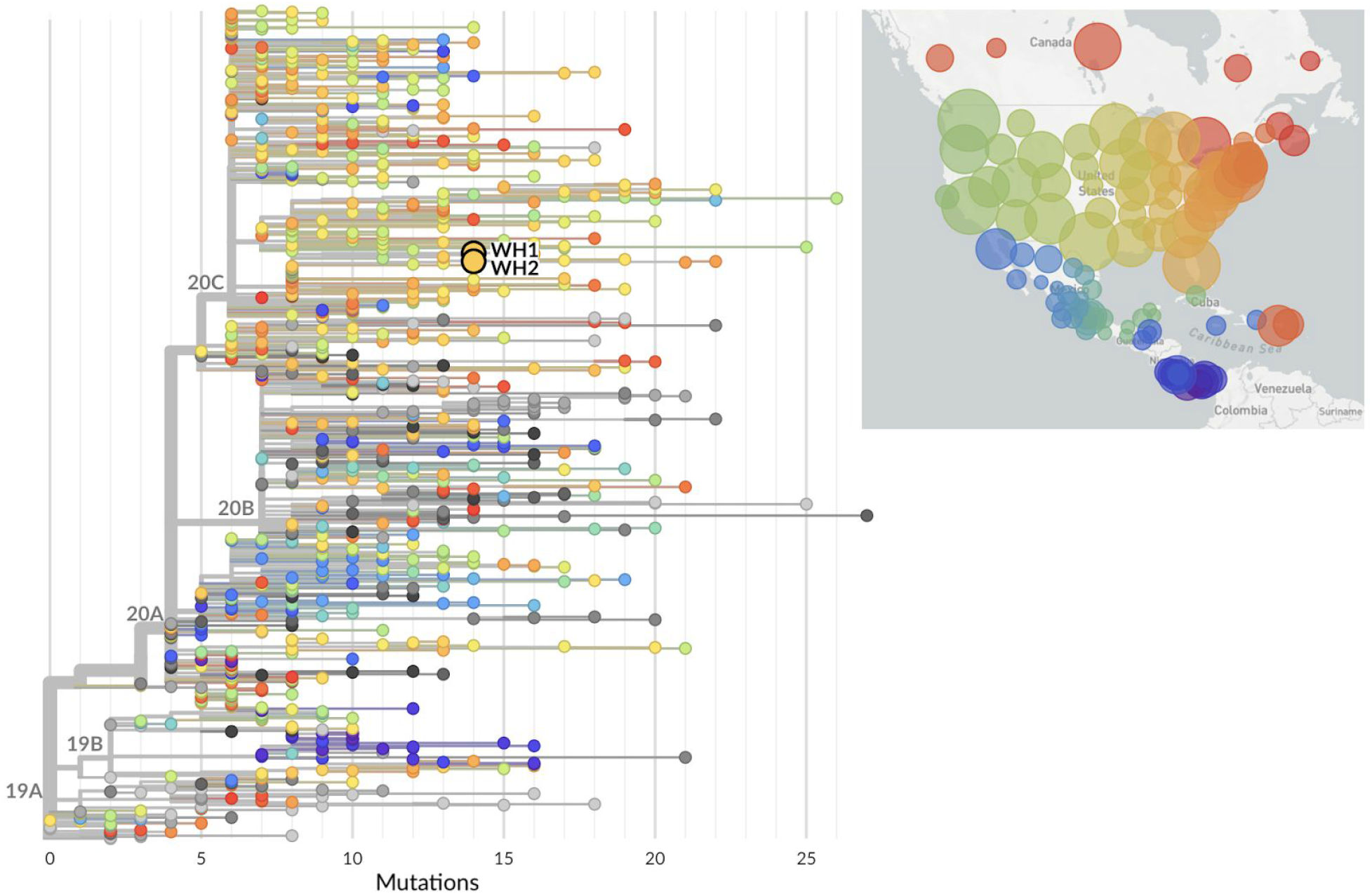
Phylogeny of 867 SARS-CoV-2 viruses collected from all over the world highlighting global placement of WH1 and WH2. Viruses from North America are preferentially sampled with 674 North American viruses (colored as shown in the inset map) and 193 viruses from outside North America (colored in gray). WH1 and WH2 are shown as larger circles with black outlines. An interactive version of this figure is available at nextstrain.org/community/blab/ncov-wh/background.

We compared WH1 and WH2 to all sequences available in the GISAID EpiCoV database [16,17] and identified viruses that are directly ancestral to WH1 and WH2 in a maximum-likelihood phylogeny (**Fig. 3A**). We find that WH1 and WH2 are descended from viruses sampled from the USA (Connecticut, Florida, New York, Texas, Washington), Canada and New Zealand in March and April 2020 with the addition of mutations A1977G, G7936T, G14250T, T18417C, C19524T and C20402T. There are related viruses that share C20402T that were sampled in Virginia in August and October 2020, but these possess other distinct mutations and consequently a molecular clock analysis places the common ancestor of WH1/WH2 and this sister clade between April and July 2020 (median estimate Apr 5, 95% confidence interval: Mar 26 to Jul 27) (**Fig. 3B**). More closely related to WH1 and WH2 is a single sample collected from Virginia in August 2020 that additionally shares A1977G with WH1 and WH2. And still more closely related are three viruses collected from Michigan in October 2020 that additionally share G7936T, G14250T and T18417C. The three Michigan viruses possess a single differentiating mutation (C27610T), while WH1 and WH2 also possess a single differentiating mutation (C19524T). A molecular clock analysis places the common ancestor between these lineages in August or September 2020 (median estimate Sep 26, 95% confidence interval: Aug 12 to Sep 30).

**Figure 3.**
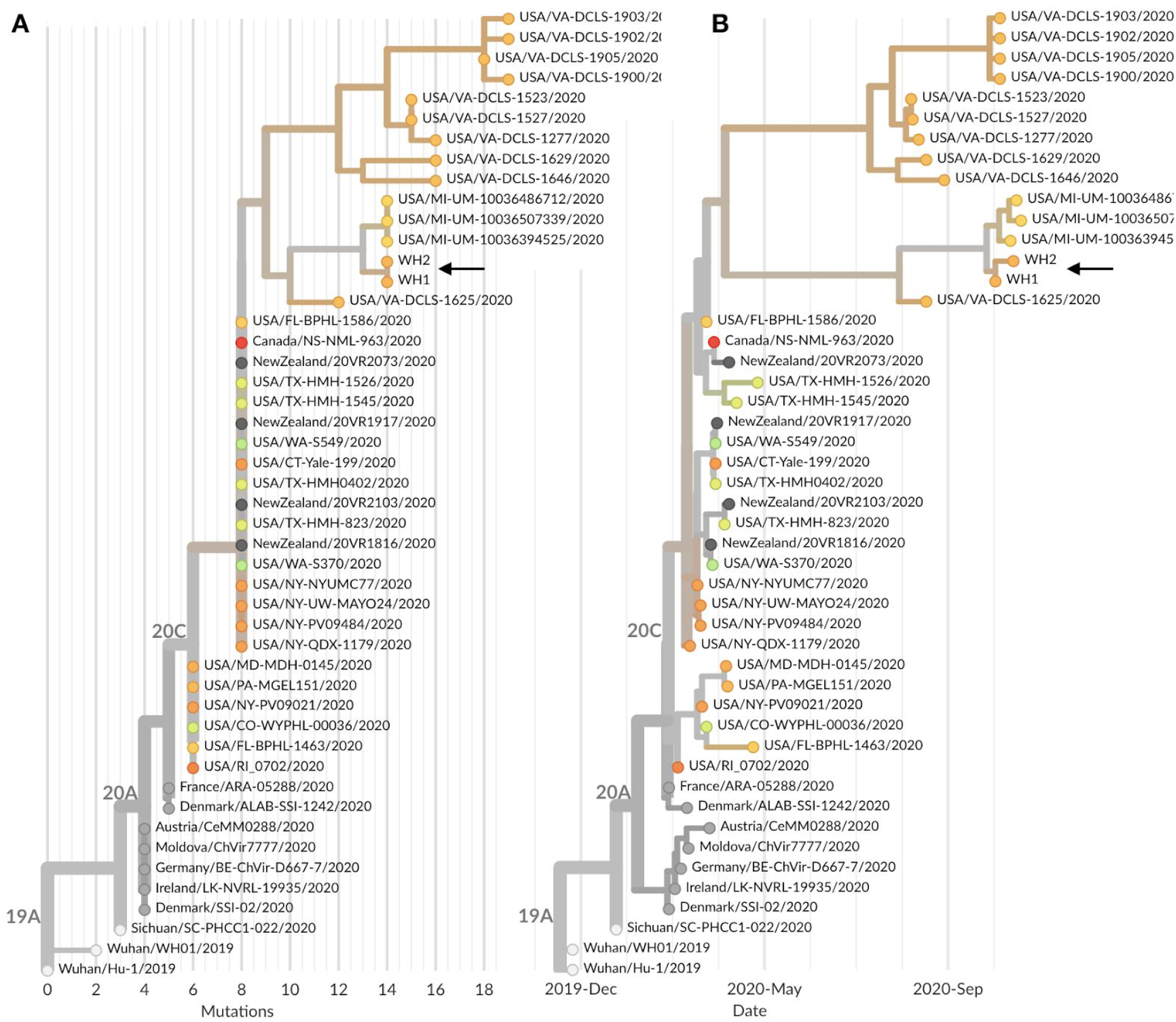
Phylogeny of 48 SARS-CoV-2 viruses that are either sister lineages to WH1 and WH2 or directly ancestral in the global maximum-likelihood phylogeny. Shown are both (A) phylogeny with branch lengths scaled by number of mutations from Wuhan reference genome and (B) temporally resolved phylogeny with branch lengths estimated according to a molecular clock analysis. Both panels are colored according to state of sampling for US samples or colored gray if samples were from outside the US. An interactive version of this figure is available at nextstrain.org/community/blab/ncov-wh/lineage.

These observations suggest a transmission chain leading to WH1 and WH2 from viruses circulating in the USA in March and April that collects an additional 6 mutations over these 6 months of circulation, consistent with the overall observed rate of molecular evolution of SARS-CoV-2 and natural Poisson variation [9]. Phylogenetic grouping suggests that this lineage may have been circulating in Virginia during this time. Notably there are 267 viruses collected between August and November 2020 from Virgina, 113 viruses from Maryland and 0 from the District of Columbia. If this lineage was circulating concurrently in DC it would not be picked up with existing sampling. The closest related viruses sampled from Michigan in October may represent a recent geographic offshoot or may represent direct geographic ancestry of the WH lineage.

## Discussion

Viral genome sequencing represents a powerful new tool for epidemiological investigation. However, it generally requires a decent fraction of infections to be sequenced to provide critical comparisons for the sequence or group of sequences in question. Nonetheless, there are situations where prompt genome sequencing of even a handful of viral genomes can be highly informative. For example, in late February 2020, genome sequencing of early SARS-CoV-2 infections in the Washington State area by us and colleagues strongly suggested cryptic spread of COVID-19 during January and February 2020, before active community surveillance was implemented [9].

The USA and the world have sequenced and publicly shared SARS-CoV-2 genomes more rapidly and at a far larger scale than any previous outbreak, epidemic or pandemic. To date, over 40,000 SARS-CoV-2 virus genomes have been sequenced and publicly shared from the USA alone [16,17]. These publicly available genomes place WH1 and WH2 along a lineage circulating (at least) in Virginia and Michigan.

Given their identical genetic sequences, we believe that WH1 and WH2 are closely related epidemiologically. Given that the individuals in question did not have any direct contact with one another and both attested exposure at events associated with the White House COVID-19 outbreak, we believe that a shared epidemiological connection through the White House COVID-19 outbreak is the most parsimonious explanation for their infections’ genetic similarity. This would imply that the WH lineage identified here was responsible for other infections in the White House cohort as well. However, we cannot completely rule out that the WH lineage identified here was circulating more broadly in the DC area and both individuals independently acquired infections of this lineage outside of White House associated exposure. Further sequencing of infections from DC and surroundings from this time period would help to definitively rule out this possibility. Sequencing of additional infections in the White House cohort would be helpful in this regard as well.

There are currently no other viruses in the GISAID EpiCoV database collected from Washington DC after August 1, 2020. Retrospective sequencing of samples from Washington DC in this time period could yield other viruses that are part of the same lineage, but this is far from certain, as the WH lineage may have been introduced from elsewhere in the US. Generally, there continues to be substantial backfill of publicly available sequences and it is possible that more viruses related to WH1 and WH2 will be sequenced and shared in the coming months. In just the period from November 1 to November 12, 10 additional sequences shared to GISAID have possessed the mutation C20402T and cluster alongside the WH lineage.

We believe that the WH lineage is likely to be at least relatively rare given its paucity of sampling in publicly available sequences. Looking forward, this relative rarity should make it possible to identify infections that likely descend from the White House outbreak. If viruses from November onwards are discovered that possess the same constellation of mutations as WH1 and WH2, then the inference would be that these infections are the downstream repercussions of the sizable October White House outbreak. This has precedent in the downstream impact of a superspreader event at a business conference in Boston in February, where conference-associated mutations were later seen at high frequency in the downstream Massachusetts epidemic [18].

The COVID-19 pandemic has damaged health and health systems and disrupted society globally to a degree and with a speed unprecedented since the 1918 influenza pandemic. Science has made a great many discoveries and innovations since then, with genome sequencing being a fairly recent addition to the toolkit to combat infectious disease. We, as a society, have the tools to control COVID-19, they just have to be employed.

## Methods

### Sample collection

Individuals were enrolled as part of the HAARVI study. All participants completed informed consent. Previously collected samples, as well as prospectively collected samples, were used for this analysis. This study was approved by the University of Washington IRB (protocol #STUDY00000959).

Specimens were shipped to the Brotman Baty Institute for Precision Medicine via commercial couriers or the US Postal Service at ambient temperatures and opened in a class II biological safety cabinet in a biosafety level-2 laboratory. Dry swabs were rehydrated with 1 mL low TE, agitated for a minute, and allowed to incubate for 10 minutes at room temperature. Two 400 µL aliquots of low TE were collected from each specimen and stored at 4°C until the time of nucleic acid extraction, performed with the MagnaPure 96 small volume total nucleic acids kit (Roche) or MagMAX Viral Pathogen II Nucleic Acid Isolation Kit (ThermoFisher). SARS-CoV-2 detection was performed using real-time RT-PCR with a probe sets targeting Orf1b and S with a FAM fluor (Life Technologies 4332079 assays # APGZJKF and APXGVC4APX) multiplexed with an RNaseP probe set with a HEX fluor (Life Technologies A30064 or IDT custom) each in duplicate on a QuantStudio 6 instrument (Applied Biosystems).

### Sequencing

SARS-CoV-2 genome sequencing was conducted using a hybrid-capture approach. RNA was fragmented and converted to cDNA using random hexamers and reverse transcriptase (Superscript IV, Thermo) and a sequencing library was constructed using the Illumina TruSeq RNA Library Prep for Enrichment kit. For hybrid capture, we used the Ct value as a proxy for viral load to balance the samples and pooled 24-plex with Seattle-based samples for the hybrid capture reaction. Capture pools were incubated overnight with probes targeting the Wuhan-Hu-1 isolate, synthesized by Twist Biosciences. The manufacturer’s protocol was followed for the hybrid capture reaction and target enrichment washes. The hybrid capture pools were sequenced on the Illumina MiSeq instrument using 2×150bp reads. The resulting reads were assembled against the SARS-CoV-2 reference genome Wuhan-Hu-1/2019 (Genbank accession MN908947) using the bioinformatics pipeline github.com/seattleflu/assembly. The consensus genome sequence of WH1 was deposited to the GISAID EpiCoV database as strain USA/DC-BBI1/2020 with accession EPI_ISL_603248 and the consensus genome sequence of WH2 was deposited to the GISAID EpiCoV database as strain USA/DC-BBI2/2020 with accession EPI_ISL_605791. Consensus genome sequences for both WH1 and WH2 are also available from github.com/blab/ncov-wh.

### Analysis

Consensus genome sequences of WH1 and WH2 were combined with SARS-CoV-2 genomes downloaded from GISAID [16,17] and processed using the Nextstrain [13] bioinformatics pipeline Augur to align genomes via MAFFT v7.4 [19], build maximum likelihood phylogeny via IQ-TREE v1.6 [20] and reconstruct nucleotide and amino acid changes on the ML tree. Branch lengths were temporally resolved using TreeTime v0.7.4 [21]. The resulting tree was visualized in the Nextstrain web application Auspice to view resulting inferences. Workflows to reproduce phylogenetic trees shown in **Figure 2** and **Figure 3** are available from github.com/blab/ncov-wh.

## Supporting information

GISAID Acknowledgements Table

## Data Availability

Consensus genome sequences and analysis code are available from https://github.com/blab/ncov-wh.

https://www.gisaid.org/

## Acknowledgements

We gratefully acknowledge the authors, originating and submitting laboratories of the sequences from GISAID’s EpiFlu Database on which this research is based. In particular we thank Virginia Division of Consolidated Laboratory Services, the University of Michigan Clinical Microbiology Laboratory and the Lauring Lab at the University of Michigan for sharing closely related samples. A full Acknowledgments table is available as supplementary materials. This work was supported by funding from the Brotman Baty Institute for Precision Medicine.

## Competing interests

HYC is a consultant for Merck and GlaxoSmithKline. JS is a consultant with Guardant Health, Maze Therapeutics, Camp4 Therapeutics, Nanostring, Phase Genomics, Adaptive Biotechnologies, and Stratos Genomics, and he has a research collaboration with Illumina. All other authors declare no competing interests.

## Notes

### Summary of Updates

Manuscript updated with full sequencing of WH2 and with the inclusion of additional sequences shared to GISAID.

